# Microstructure predicts non-motor outcomes following Deep Brain Stimulation in Parkinson’s disease

**DOI:** 10.1101/2023.04.25.23289088

**Authors:** Philipp A. Loehrer, Miriam H. A. Bopp, Haidar S. Dafsari, Sieglinde Seltenreich, Susanne Knake, Christopher Nimsky, Lars Timmermann, David J. Pedrosa, Marcus Belke

**Affiliations:** Department of Neurology, Philipps-University Marburg, Marburg, Germany; Department of Neurosurgery, Philipps-University Marburg, Marburg, Germany; Center for Mind, Brain and Behavior (CMBB), Marburg, Germany; Department of Neurology, University Hospital Cologne, Cologne, Germany; Center for Personalized Translational Epilepsy Research (CePTER) Consortium

**Keywords:** non-motor symptoms, Parkinson’s disease, deep brain stimulation, diffusion imaging, NODDI, microstructure

## Abstract

**Background:** Deep brain stimulation of the subthalamic nucleus (STN-DBS) is an effective treatment for motor and non-motor symptoms in advanced Parkinson’s disease (PD). However, considerable interindividual variability of outcomes exists. Neuroimaging based biomarkers, such as neurite orientation dispersion and density imaging (NODDI), a biophysical model based MRI-technique, have been proposed to predict clinical outcomes and therefore inform preoperative patient counselling.

**Objective:** To detect microstructural properties of brain areas associated with short-term non-motor outcomes following STN-DBS in PD.

**Methods:** In this prospective open-label study, 37 PD patients underwent diffusion MRI and comprehensive clinical assessments at preoperative baseline and 6-month follow-up. Neurite density index (NDI), orientation dispersion index (ODI), and fractional anisotropy (FA) were derived. Whole brain voxel-wise analysis assessed associations between microstructural metrics and non-motor outcomes corrected for multiple comparisons using a permutation-based approach.

**Results:** Intact microstructure within specific areas including right insular cortex, right putamen, right cingulum, and bilateral corticospinal tract were associated with greater postoperative improvement of non-motor symptom burden. Furthermore, microstructural properties of distinct brain regions were associated with postoperative changes in sleep, attention/memory, and urinary symptoms.

**Conclusion:** Microstructural properties of distinct brain areas predict non-motor outcomes in DBS for PD. Therefore, diffusion MRI can support preoperative patient counselling and treatment selection by identifying patients with above-or below-average non-motor responses.

## Introduction

Deep brain stimulation (DBS) of the subthalamic nucleus (STN) is an established therapy for advanced Parkinson’s disease (PD), improving motor- and non-motor symptoms.^1-3^ Despite significant therapeutic benefits at the group level, there is large variability in outcomes at the individual subject-level, with some patients even experiencing persistent symptoms.^4^ To predict postoperative outcomes and thereby improve preoperative patient counselling, the use of neuroimaging-based markers has been proposed.^4^ In this regard, quantitative MRI techniques such as neurite orientation dispersion and density imaging (NODDI) have gained increasing attention in recent years to track disease progression and treatment response.^5^ NODDI is a multi-compartmental diffusion-weighted MRI technique which facilitates the assessment of specific microstructural properties directly related to neurite morphology.^6^ The model provides two voxelwise metrics of neurite morphology: the neurite density index (NDI), describing the density of axons and dendrites within a voxel, and the neurite orientation dispersion index (ODI), characterizing the variability of neurite orientations, i.e. how parallel they are. Importantly, the relationship between NODDI metrics and underlying tissue properties has recently been confirmed histologically.^7^ Despite its sensitivity and specificity, NODDI protocols have a clinically feasible data acquisition time, making them an important tool for use in clinical research.^6^ Previous studies employing NODDI in PD have shown that the model is capable of differentiating PD patients from healthy controls^11^ and patients with atypical Parkinsonism.^12^ Furthermore, NODDI characterized disease related pathology such as retrograde degeneration of the nigrostriatal pathway,^13^ and its metrics were associated with bimanual motor control,^14^ disease severity, and duration.^6,11^ Combining NODDI with conventional DTI metrics in a multi-parametric analysis therefore provides complementary information on microstructural properties and may serve as an imaging-based marker to facilitate treatment prediction and inform patient counselling. Thus, we sought to demonstrate that a multi-parametric quantitative MRI approach can detect microstructural properties of brain areas that predict non-motor outcomes following STN-DBS in PD. Specifically, we aimed to identify regions whose microstructural metrics, i.e. NDI, ODI, and fractional anisotropy (FA), were associated with (1) changes in overall non-motor symptom burden and (2) changes in non-motor symptom domains, showing significant improvements after STN-DBS. The results of the present study should help to guide preoperative patient counselling by identifying microstructure that predicts above-or below-average non-motor response to STN-DBS.

## Methods

The study was approved by the local ethics committee (study-number: 155/17) and carried out in accordance with the Declaration of Helsinki.

### Participants

Thirty-seven PD patients (9 female, mean age 58.8 ± 7.3 years) were enrolled in this prospective, observational, ongoing study upon written informed consent (for demographics cf. Table 1). Inclusion criteria comprised indication for DBS lead surgery because of advanced PD according to modern criteria.^8^ Patients were excluded if they had pathological MR imaging, a concomitant neurological or psychiatric disease, or impaired visual or auditory function.

**Table 1:** Baseline characteristics and outcomes at baseline and 6-month follow-up Demographic characteristics and outcome parameters at baseline and 6-months follow-up. Reported p-values are corrected for multiple comparisons using Benjamini-Hochberg’s method. Bold font highlights significant results, p<.05. **Abbreviations:** 6-MFU = 6-month follow-up; LEDD = Levodopa equivalent daily dose; LEDD-DA: LEDD of Dopamine Agonists; NMSS = Non-Motor Symptom Scale; PDQ-8 SI = 8-item Parkinson’s Disease Questionnaire summary index; SCOPA = Scales for Outcomes in Parkinson’s disease;

|  | *n* | *M* | *SD* |  |  |  |  |  |
| --- | --- | --- | --- | --- | --- | --- | --- | --- |
| Age [y] | 37 | 58.8 | 7.3 |  |  |  |  |  |
| Disease duration [y] | 37 | 8.9 | 4.3 |  |  |  |  |  |
| Sex (female/male) [%] | 37 | 9/28 | [24.3/75.7] |  |  |  |  |  |
|  | Baseline |  |  | 6-MFU |  |  | Baseline vs.<br>6-MFU |  |
|  | *n* | *M* | *SD* | *n* | *M* | *SD* | *p* | *effect size* |
| NMSS total score | 37 | 71.9 | 39.1 | 37 | 48.4 | 29.1 | **.007** | **-.34** |
| Cardiovascular | 37 | 1.6 | 2.7 | 37 | 1.1 | 2.5 | .225 | -.15 |
| Sleep/fatigue | 37 | 16.9 | 11.5 | 37 | 7.9 | 7.6 | **<.001** | **-.44** |
| Mood/ apathy | 37 | 11.6 | 13.3 | 37 | 6.0 | 7.6 | .109 | -.21 |
| Perceptual problems/<br>hallucinations | 37 | .9 | 2.5 | 37 | 1.1 | 3.0 | .865 | -.03 |
| Attention/ memory | 37 | 7.0 | 6.5 | 37 | 5.0 | 5.9 | **.022** | **-.29** |
| Gastrointestinal | 37 | 5.6 | 5.0 | 37 | 4.4 | 4.9 | .223 | -.16 |
| Urinary | 37 | 13.2 | 10.4 | 37 | 9.1 | 10.2 | **.026** | **-.28** |
| Sexual function | 37 | 3.8 | 5.0 | 37 | 3.0 | 4.3 | .296 | -.13 |
| Miscellaneous | 37 | 11.2 | 8.9 | 37 | 10.7 | 7.8 | .865 | -.02 |
| PDQ-8 SI | 37 | 32.3 | 14.8 | 37 | 22.6 | 14.1 | **<.001** | **.67** |
| SCOPA-M total score | 37 | 19.8 | 6.2 | 37 | 12.9 | 6.4 | **<.001** | **-.47** |
| SCOPA-M-motor<br>examination | 37 | 9.4 | 4.2 | 37 | 6.7 | 4.1 | **.008** | **-.33** |
| SCOPA-M-activities of<br>daily living | 37 | 6.6 | 2.7 | 37 | 4.1 | 3.1 | **.003** | **-.38** |
| SCOPA-M-motor<br>complications | 37 | 3.8 | 2.3 | 37 | 2.1 | 2.4 | **<.001** | **-.44** |
| LEDD [mg] | 37 | 962.3 | 393.2 | 37 | 537.6 | 269.0 | **<.001** | **1.2** |
| LEDD DA [mg] | 37 | 261.8 | 133.6 | 37 | 170.6 | 123.8 | **<.001** | **.7** |

### Clinical Assessment

Patients were assessed preoperatively and six months after DBS lead surgery, with medication at both times and DBS switched on postoperatively. All subjects underwent a neuropsychological assessment including the Non-Motor Symptom Scale (NMSS), the Parkinson’s Disease Questionnaire (PDQ)-8 and the Scales for Outcomes in PD -Motor Function (SCOPA-M) using standardized case report forms. Detailed information on the clinical assessment is provided in the supplementary material.

### MRI Data Acquisition and Processing

PD patients were scanned at baseline on a 3-Tesla Trio scanner (Siemens, Erlangen, Germany) at the Core Unit Brain Imaging of the University of Marburg. The acquisition protocol is reported in the supplementary material. All images were investigated to be free of motion or ghosting and high frequency and/or wrap-around artefacts at the time of image acquisition.

### Image Processing

Image analysis was performed within the FreeSurfer image analysis suite 7.1.1 (http://surfer.nmr.mgh.harvard.edu) as reported previously by our group.^9^ The processing of T1-weighted scans included skull stripping, automated Talairach transformation, cortical and subcortical segmentation, intensity normalisation, tessellation of the grey/white matter boundary, automated topology correction, and surface deformation following intensity gradients.^10^

DTI data were processed using FMRIB Software Library (FSL) 6.0.5.2 (https://fsl.fmrib.ox.ac.uk/fsl). To correct for eddy-current distortions and involuntary movements, raw DTI volumes were linearly registered and resampled to the first b0 volume.^11^ Subsequently, the diffusion tensor for each voxel was fit to the data using linear regression and FA was derived from the diffusion tensor.^12^ Furthermore, NODDI-DTI,^13^ a modification of NODDI,^6^lJ was used to obtain NDI and ODI from the DTI data. Visual inspection of the b0-images confirmed that no changes beyond those in the tissue structure contributed to the observed effects. To perform regional analyses, the first b0 image of each scan was linearly registered to the structural T1-weighted image using a boundary based method, yielding an affine matrix.^14^ Subsequently, T1-derived segmentations and brain masks were transformed to the diffusion space via the inverse of the affine matrix.

### Statistical analysis

Statistical analysis of clinical outcomes was performed in MATLAB (The MathWorks, Inc., R2018a). Changes between baseline and follow-up were analysed using the Wilcoxon signed-rank or t-test when parametric test criteria were met. The type-I error was controlled using the Benjamini-Hochberg method and effect sizes were calculated according to Cohen. Relationships between change scores of clinical data and NMSS total score (NMSS-T) were explored using Spearman correlations as reported previously.^15,16^

Statistical voxelwise analysis of image data was performed using a generalized linear model. First, FA-maps were co-registered to the MNI152 space using linear and nonlinear transformation.^17^ Masked FA-, NDI-, and ODI-maps were subsequently registered to the MNI152 space using the transformation from the previous step. Only voxels of brain tissue existing in every subject were included in the analysis. Significant associations between metrics of microstructure and change of non-motor symptoms were carried out for the whole brain as described previously.^18^ Here, percentage differences between baseline and follow-up values were calculated for NMSS-T and NMSS domains with significant postoperative improvement ((2) sleep/fatigue, (5) attention/memory, and (7) urinary). A permutation-based approach based on the Analysis of Functional NeuroImages (AFNI) null-z simulator was used to corrected for multiple comparisons employing 12,000 simulations under the null hypothesis.^19^ Clusters were formed using a threshold of p<.01 and a clusterwise p-value was calculated. Results were accepted as significant with clusterwise p<.05.

## Results

### Clinical outcomes

Longitudinal changes of clinical outcomes are reported in Table 1. At the 6-month follow-up, we observed improvements of NMSS-T (z=-2.95, p=.007, r=-.34), PDQ-8 SI (t(36)=4.22, p<.001, Cohen’s d: .67), and SCOPA-M total score (z=-4.01, p<.001, r=-.47), as well as reductions of LEDD (t(36)=7.63, p<.001, Cohen’s d: 1.2) and LEDD-DA (t(36)=4.98, p<.001, Cohen’s d: .7). Analysis of NMSS domains revealed beneficial effects of STN DBS on sleep/fatigue (domain 2; z=-3.79, p<.001, r=-.44), attention/memory (domain 5; z=-2.49, p=.022, r=-.29), and urinary symptoms (domain 7; z=-2.39, p=.026, r=-.28). Analysis of SCOPA-M domains showed improvements in motor examination (z=-2.87, p=.008, r=-.33), activities of daily living (z=-3.27, p=.003, r=-.38), and motor complications (z=-3.76, p<.001, r=-.44) at the 6-month follow-up. Results of the correlation analysis are reported in the supplementary materials (Table e-1).

### Interaction between Fractional Anisotropy and postoperative non-motor symptom change

Higher FA-values in the right insular cortex were associated with greater postoperative NMSS-T reduction (positive cluster P1, cluster wise p-value (CWP): .032). Furthermore, lower FA values were found in clusters including the bilateral cingulum and the left inferior longitudinal fasciculus (ILF), which were related to greater postoperative NMSS-T reduction (negative clusters N1-3, CWP: <.001-.014). Regional mean fractional anisotropy values associated with postoperative change in non-motor symptom burden are detailed in Table e-2 and Figures e-1 and e-2.

### Interaction between NODDI-parameters and postoperative non-motor symptom change

Whole brain analysis of NODDI parameters showed that both, ODI and NDI, were associated with changes in postoperative NMSS-T. Higher ODI-values in the right putamen, the right cingulate cortex, and the left forceps major were related to a higher postoperative NMSS-T reduction (P1-4, CWP: <.001-.034, Figure 1, Table e-3). Furthermore, lower ODI-values in regions of both corticospinal tracts as well as left occipital fusiform gyrus were related to a higher postoperative NMSS-T reduction (N1-4, CWP: <.001-.047, Figure 2, Table e-3). Lower NDI-values in the left postcentral gyrus, left cingulum, and right forceps minor were associated with a higher postoperative NMSS-T reduction (N1-4, CWP: <.001-.047, Figure e-4, Table e-4). No positive associations between NDI-values and postoperative NMSS-T change were detected.

**Figure 1.**
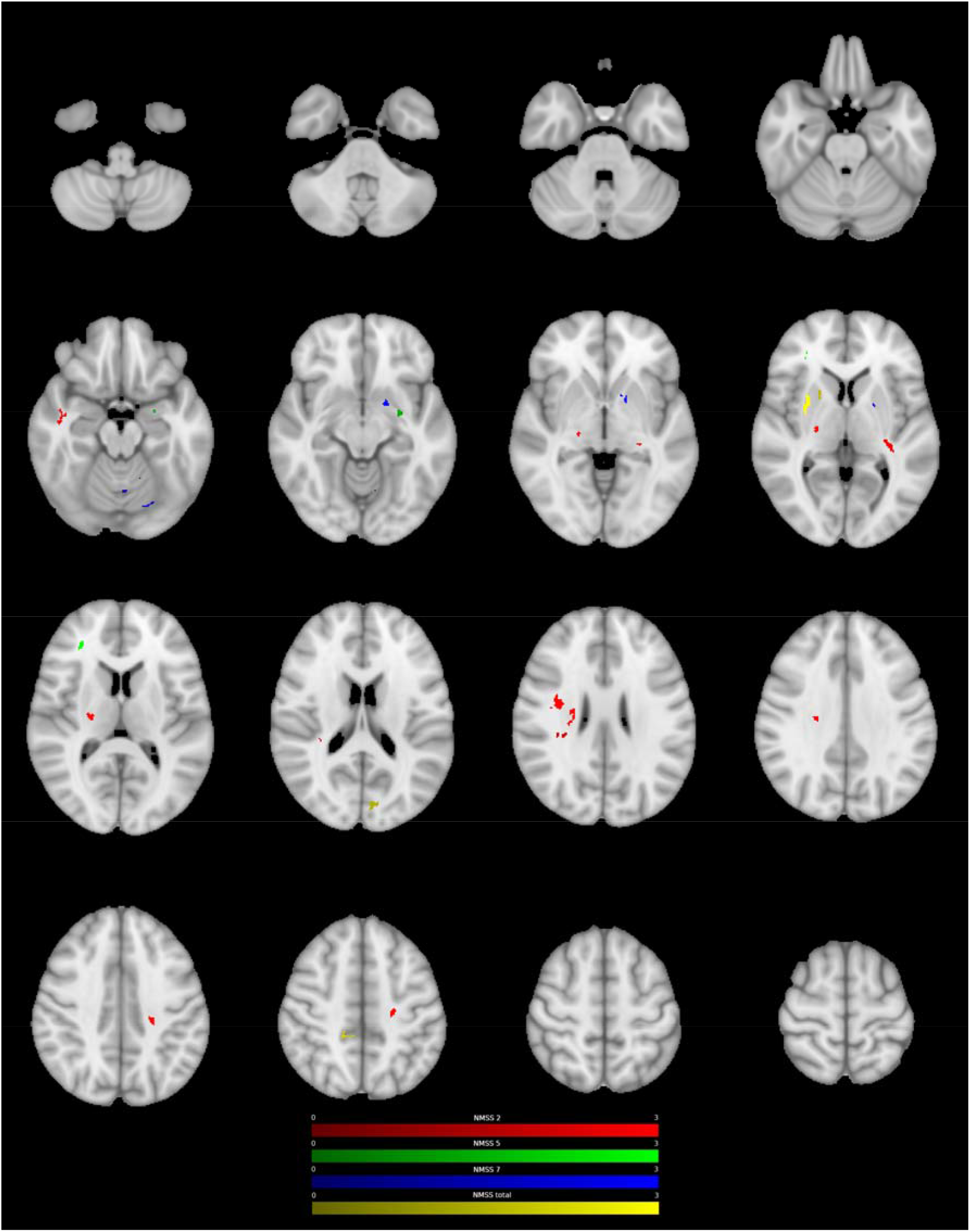
Clusters with a positive association between PD patients’ ODI-values and postoperative change in NMSS-T (yellow), Domain 2 (sleep/fatigue, red), Domain 5 (attention/memory, green), and Domain 7 (urinary, blue), as revealed by the whole brain analysis. P-Values were corrected for multiple comparisons using a permutation-based approach.

**Figure 2.**
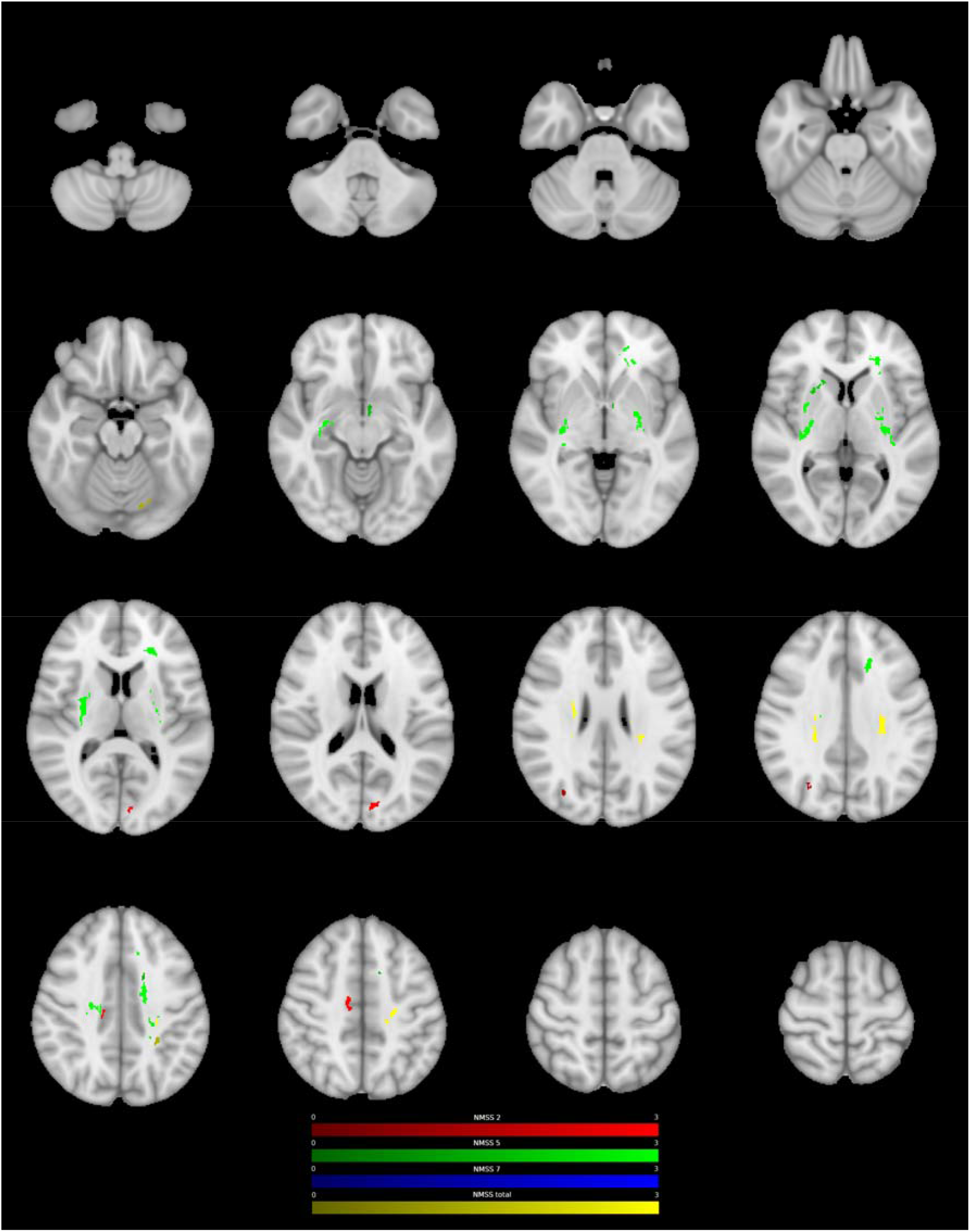
Clusters with a negative association between PD patients’ ODI-values and postoperative change in NMSS-T (yellow), Domain 2 (sleep/fatigue, red), Domain 5 (attention/memory, green), and Domain 7 (urinary, blue), as revealed by the whole brain analysis. P-Values were corrected for multiple comparisons using a permutation-based approach.

### Interaction between microstructure and postoperative change in Non-Motor Symptoms Scale domains

Regional mean values of microstructural metrics associated with significant postoperative changes in non-motor symptoms scale domains are detailed in Table e-5 (sleep/fatigue), Table e-6 (attention/memory), and Table e-7 (urinary symptoms). Higher values of microstructural metrics within vast regions of bilateral corticospinal tract (CST) were related to a higher symptom reduction in the sleep and fatigue domain (FA: P1-2, CWP: .001-.005; ODI: P1-4, CWP: <.001; NDI: P1, CWP: .004; Table e-5). With regard to attention and memory, higher FA-values in bilateral cingulum, left insular cortex, and left anterior thalamic radiation (AThR) were associated with higher postoperative symptom reduction (P1-4, CWP: <.001-.031, Table e-6). Furthermore, higher ODI-values in left parahippocampal gyrus and within the right frontal pole were related to positive postoperative outcomes in attention and memory (P1-2, CWP: <.001-.049), whereas lower ODI-values in bilateral AThR and left cingulum were associated with higher symptom reduction (N3, N5, N8-10, CWP: <.001-.039). Also, vast areas of bilateral CST and putamen that had lower ODI- and NDI-values were associated with higher symptom reduction in the attention/memory domain (ODI: N1, N2, N4 and N7-8, CWP: <.001-.011; NDI: N1-6 CWP: <.001-.023). Concerning urinary symptoms, higher ODI- and NDI-values in left putamen, pallidum, and AThR as well as left cerebellar lobule V and VI were related to higher postoperative symptom reduction in the urinary domain (ODI: P1-3, CWP: <.001-.031; NDI: P1-2, P4, CWP: <.001-.002, Table e-7). This association was also present between NDI-values and right pallidum and putamen (P3, CWP: .002). Furthermore, higher FA-values in right cingulate gyrus and left superior longitudinal fasciculus (SLF) were associated with higher postoperative reduction of urinary symptoms (P1-2, CWP: <.001-.01).

## Discussion

In the present study, NODDI-DTI, a novel method for analysing DWI data, was applied to preoperative imaging to investigate cerebral microstructure associated with non-motor symptom changes following neurostimulation in PD. There are two key findings. First, we demonstrate that intact microstructure within specific areas including right insular cortex, right putamen, right cingulum, and bilateral CST were associated with higher reduction of postoperative non-motor symptom burden. Second, we delineate the structures and their microstructural properties which are associated with postoperative improvements in specific non-motor domains.

Whole brain analysis of NODDI parameters identified an association between higher ODI in right putamen as well as right cingulum and a higher reduction of postoperative non-motor symptom burden. As ODI is high in gray matter,^6^ this finding supports the notion that intact sprawling of dendritic processes in these areas is important for beneficial postoperative non-motor outcomes. In PD, the pathological deposition of alpha-synuclein in intraneuronal Lewy inclusions is accompanied by a degeneration of neurons and severe morphological changes of dendrites.^20,21^ These tissue changes cannot be seen in conventional MRI, whereas NODDI is sensitive to neurite morphology.^6^ Indeed, previous work demonstrated reduced putaminal ODI-values in patients with PD compared to healthy controls.^21^ This finding was interpreted as decreased dendrite length and loss of spines of striatal medium spiny neurons, which are the primary target of dopaminergic nigrostriatal projections.^21^ Considering the close topographical relationship between STN and putamen as well as insular cortex and the integration of STN in basal ganglia-thalamo-cortical loops with its motor, associative, and limbic projections, it is important to examine the assumed mechanisms of action of DBS.^4,22^ Besides effects on the micro- and mesoscale, the high-frequency pulses of electrical current emitted by DBS electrodes affect interregional networks on the macroscale.^4^ Here, modulation of networks has been able to predict postoperative outcomes across several motor and non-motor symptoms.^4^ Furthermore, previous work has shown that compromised putaminal microstructure in PD patients can be associated with higher non-motor symptom burden independent of motor symptoms.^23^ Therefore, it can be hypothesised that the positive association between ODI and postoperative non-motor outcomes in the present study represents the dependency of DBS on intact tissue structure to exert its network effects.

Besides associations with gray matter areas, ODI showed a negative association with postoperative non-motor symptom burden in bilateral CST. In healthy white matter tissue, ODI is usually low, as fibers are highly coherent to another whereby high values of ODI represent axonal disorganisation and degeneration.^6^ Therefore, the observed negative association might reflect the dependence of DBS on intact tissue structure in CST, as low ODI, i.e. intact white matter microstructure, was associated with beneficial postoperative outcomes. Previous DTI-studies have repeatedly demonstrated microstructural alterations of CST in PD. In particular, increased FA has consistently been reported and hypothesised to demonstrate a compensatory mechanism, reflecting axonal sprouting secondary to a reduced input from striatum and thalamus.^24^ When, however, microstructure in CST deteriorates over the course of the disease, an association with motor dysfunction was demonstrated.^25^ Associations between altered CST-microstructure and non-motor symptoms, on the other hand, have received little attention. Employing a connectometry analysis in 85 patients, Ashraf-Ganjouei and colleagues could demonstrate that lower axonal density in CST was associated with a higher burden of gastrointestinal symptoms.^26^ Furthermore, lower FA-values in right CST were observed in PD patients with depression compared to non-depressed patients.^27^ Extending these findings, the results of the present study demonstrate that not only non-motor symptoms but also beneficial non-motor outcomes following STN-DBS depend on intact CST microstructure. Considering the role of the CST as major effector of motor control, motor outcomes and the postoperative reduction in dopaminergic medication could contribute to the beneficial non-motor effects in the present study. Importantly, however, postoperative improvements in non-motor symptom burden were not related to improvements in LEDD, LEDD-DA, and SCOPA-motor examination scores, which is in accordance with the literature.^28,29^ Therefore, the association between intact CST microstructure and beneficial postoperative non-motor outcomes seems to be independent of the motor effects of DBS and suggests that CST microstructure is inherently relevant for non-motor symptoms in PD.

### Microstructure is associated with beneficial sleep outcomes

Sleep disturbances affect the majority of PD patients and result in poor quality of life.^30^ Associated disorders encompass both, disturbances of sleep-wake transition, as well as parasomnias.^30^ Although the exact neural mechanisms remain to be established, a disrupted interaction of neuronal circuits and different neurotransmitter systems has been suggested to underlie the sleep disturbances in PD.^31,32^ STN-DBS has been suggested to improve sleep by alleviating motor symptoms and directly altering sleep physiology resulting in increased total sleep time, sleep efficiency, and quality of sleep as well as reduced wakefulness after sleep onset and insomnia.^30^ In the present study, STN-DBS improved symptoms of the sleep/fatigue domain and beneficial outcomes were associated with higher FA-, ODI-, and NDI-values in vast regions of the CST. On the other hand, lower ODI- and NDI-values in these areas were associated with detrimental or below average response to STN-DBS. As most of the clusters were overlapping and within regions of high fiber crossing and dispersion, the selective degeneration of crossing fibers might underlie the observed relationship. The results, therefore, support the hypothesis that intact microstructure of the CST and its crossing fibers is important for beneficial postoperative changes in the sleep and fatigue domain. Additionally, high ODI-values of cortical structures including right superior and middle temporal gyrus and right parietal operculum cortex were associated with beneficial postoperative outcomes. Previous studies of healthy controls using simultaneous recordings of electroencephalography and functional MRI during sleep reported an association of activity increases within these areas with sleep spindles during early stages of non-rapid eye movement (NREM) sleep.^33^ In PD, spindle density and amplitude seems to be reduced during NREM and modulated by dopaminergic therapy.^30^ Integrating these findings with results of the present study one could speculate that intact microstructure, i.e. dendritic arborisation, in these areas is important for STN-DBS to modulate sleep physiology.

### Microstructure is associated with beneficial attention and memory outcomes

Higher FA (left hemisphere) and lower ODI in bilateral AThR were associated with beneficial postoperative outcomes in the attention and memory domain. These results suggest, that degenerative changes in axonal structure underlying FA alterations in AThR are attributable to changes in axonal fanning and dispersion. AThR connects the frontal lobe, the dorsolateral prefrontal cortex (DLPFC) in particular, with the anterior and midline nuclei of the thalamus.^22^ As these nuclei are integrated in functional loops with the cingulum and the pallidum, the nuclei and their fiber connections are associated with the limbic system and thought to be involved in executive functions and planning of complex behaviour.^22,34^ The DLPFC is involved in various higher-level cognitive functions including attention, working memory, and executive control.^35,36^ Considering the functional association of AThR with STN, as well as its role in connecting the structures named above, it seems reasonable that intact microstructure in AThR is important for the conveyance of beneficial effects on memory function and attention.

Higher FA and lower ODI in overlapping clusters in bilateral cingulum as well as associated structures were related to increases in postoperative attention and memory function, indicating that axonal fanning and dispersion underlies alterations in FA in the cingulum. The cingulum is a group of nerve fibers connecting the hippocampus, prefrontal, parietal, and anterior cingulate cortex.^37^ This allows for the integration of information from these structures and explains the involvement of the cingulum in attention, memory, and emotion regulation.^37^ Previous studies in PD have shown that compromised microstructure of the cingulum was related to reduced scores in cognitive assessments, impaired visuospatial memory, and dementia.^38^ Taken together, the association between microstructural properties and beneficial attention and memory outcomes in bilateral AThR and cingulum might represent the dependency of DBS on intact tissue structure to exert its network effects.

### Microstructure is associated with beneficial outcomes of urinary symptoms

Deficient perception of multimodal sensory information is a characteristic of PD leading to debilitating non-motor symptoms.^39^ Sensory deficiencies in PD have been described in both, somatosensory pathways associated with proprioception as well as visceral pathways involved in monitoring of urinary bladder filling.^40^ STN-DBS in PD was shown to improve the perception of urinary bladder filling, resulting in a delayed desire to void and increased bladder capacity.^39^ Improved urinary function was attributed to a beneficial influence of STN-DBS on a gain of afferent bladder information due to an increase or decrease of activation of primary sensory areas.^39^ In particular, previous studies showed an activation of anterior cingulate gyrus (ACC) and left lateral frontal cortex during monitoring and controlling the storage phase of the urinary cycle and these structures are thought to be involved in the urge to void, withholding urine, and the onset of micturition.^39^ In the present study, intact microstructure, i.e. high FA, in right ACC was associated with postoperative improvements in urinary symptoms suggesting that sound microstructure in ACC is necessary for STN-DBS to have beneficial effects on controlling the storage phase of the urinary cycle. Furthermore, high NDI and ODI, i.e. high dispersion without a decrease in axonal density, in left AThR was associated with positive outcomes of urinary symptoms. AThR connects the frontal cortex with the thalamus and the pallidum and previous studies suggested that modulation of left frontal cortex by STN-DBS is important for urge control.^39^ Therefore, intact microstructure in left AThR might be relevant for STN-DBS to modulate left frontal cortex during urge control.^39^ Further important structures implicated in processing afferent urinary bladder information are posterior thalamus, which is activated during bladder filling and micturition, and ventrolateral as well as reticular thalamus, which receive input from the striatum to modulate the flow of visceral information between posterior thalamus and the cortex. Previous studies speculated that STN-DBS may recondition the interaction between pallidal output and the modulatory effect of the thalamus, resulting in improved gating of sensory information.^39^ One might speculate, that the association of intact microstructure in left pallidum and putamen with better outcomes in the urinary domain represents the importance of these structures for the effect of STN-DBS on gating sensory information.

### Limitations

Three main limitations of our study have to be addressed. First, despite histopathological validation of the NODDI model and its frequent use in PD, no studies validating the model in post-mortem brain tissue of PD patients exist. Second, the underlying assumptions of the NODDI model may represent an oversimplification and might therefore result in reduced specificity.^6^ Third, the resolution of the DTI scan is limited to 2.0×2.0×2.0mm, which might be too coarse for valid assessments of small fiber bundles.

## Conclusion

In conclusion, we describe a spatially distinct profile of microstructural alterations associated with beneficial non-motor outcomes following neurostimulation in PD, including right insular cortex, putamen, cingulum, and bilateral corticospinal tract. Furthermore, we delineate the structures and their microstructural properties which are associated with postoperative improvements in specific non-motor domains. Therefore, we suggest that diffusion MRI can support preoperative patient counselling by identifying patients with above-or below-average non-motor responses.

## Data Availability

All data produced in the present study are available upon reasonable request to the authors

## Acknowledgement

The authors would like to thank the participants for their active engagement in this study.

## Data and Code Availability

The data that support the findings of this study are available on request from the corresponding author (PAL). The data are not publicly available due to privacy or ethical restrictions. All tools used for the analysis of MRI data are based on FreeSurfer Version 7.1 (http://surfer.nmr.mgh.harvard.edu/) and FSL 6.0.5.2 (http://www.fmrib.ox.ac.uk/fsl) packages, which are freely available. Scripts for automation were written in tcshell and parts of the statistics were written in Python using the packages numpy, pandas, seaborn, matplotlib, nibabel and scipy, which are also freely available. Python program code for the analysis of NODDI-DTI is available from https://github.com/dicemt/DTI-NODDI.

## Contributorship

PAL: study concept and design, data acquisition, data analysis, drafting of the manuscript

MBo: data acquisition, surgical intervention, critical revision of the manuscript

HSD: study design, critical revision of the manuscript

SS: data acquisition, critical revision of the manuscript SK: critical revision of the manuscript

CN: data acquisition, surgical intervention, critical revision of the manuscript LT: study design, critical revision of the manuscript

DJP: study concept and design, data acquisition, data analysis, drafting of the manuscript MBe: study concept and design, data acquisition, data analysis, drafting of the manuscript

