## supplementary_material for "Microstructure predicts non-motor outcomes following Deep Brain Stimulation in Parkinson’s disease"

**Supplementary Methods**

**Clinical Assessment**

Demographic and clinical data were collected on both study visits for each participant using standardized case report forms. Patients underwent a comprehensive neuropsychological assessment including the following scales:

Non-motor Symptoms Scale (NMSS): the clinician rated scale comprises 30 items evaluating 9 dimensions of non-motor symptoms including (1) cardiovascular, (2) sleep/fatigue, (3) mood/cognition, (4) perceptual problems/hallucinations, (5) attention/memory, (6) gastrointestinal tract, (7) urinary, (8) sexual function, and (9) miscellaneous which in turn asks about pain, the ability to smell/taste, weight change, and sweating. The score ranges from 0 (no impairment) to 360 (maximum impairment) and assesses the NMS over the past 4 weeks.^1^

PD Questionnaire (PDQ)-8: the questionnaire is a short form of the PDQ-39 and determines eight dimensions of quality of life (QoL) in PD patients.^2^ The score is a well-established measure in PD patients undergoing DBS surgery and reported as a summery index (SI) with a score range from 0 (no impairment) to 100 (maximum impairment).^3-5^

Scales for Outcomes in PD (SCOPA) – Motor Function: the clinician rated scale evaluates 3 dimensions of motor function in PD, comprising (A) motor evaluation, (B) activities of daily living, and (C) motor complications, with subscale scores ranging from 0 (no impairment) to 42 (motor evaluation), 21 (activities of daily living), and 12 (motor complications), respectively.^6^

Levodopa equivalent daily dose was calculated according to Tomlinson et al.^7^

**MRI Data Acquisition and Processing**

Parkinson’s disease patients in the MedON were scanned at baseline on a 3-Tesla Trio scanner (Siemens, Erlangen, Germany) at the Core Unit Brain Imaging of the University of Marburg. The acquisition protocol comprised the following sequences:

1. 3D T1-weighted Magnetization Prepared - RApid Gradient Echo sequence (MPRAGE, field of view (FoV) = 256 mm, matrix 256x256, 176 slices, slice thickness 1 mm, voxel dimension 1.0x1.0x1.0 mm³, repetition-time (TR) = 1900 ms, echo-time (TE) = 2.26 ms, inversion time (TI) = 900 ms, flip-angle = 9°, bandwidth (BW) = 200 Hz/Pixel, parallel imaging (GRAPPA) with factor 2)
2. diffusion weighted (DWI) (FoV = 256 mm, matrix 128x128, slice thickness 2 mm, distance factor 0 %, voxel dimension 2.0x2.0x2.0 mm³, TR = 7900 ms, TE = 90 ms, BW = 1502 Hz/Pixel, 42 diffusion encoding gradients, three intermittent non-weighted b0 images (b = 0 s/mm²), high b-value b = 1000 s/mm², GRAPPA with factor 2).

All images were investigated to be free of motion or ghosting and high frequency and/or wrap-around artefacts at the time of image acquisition.

**Supplementary Results**

**Correlation analysis**

**Table e-1 Correlations between change scores of clinical data and NMSS-T**

|  | NMSS-Total change score | | |
| --- | --- | --- | --- |
| Change scores | N | *p*-value | *effect size* |
| PDQ-8 Summary Index | 37 | .117 | .26 |
| SCOPA-  Total  Motor evaluation  Activities of daily living  Motor complications | 37  37  37  37 | **.037**  .288  **.009**  .495 | **.34**  .18  **.43**  .16 |
| LEDD total  LEDD-DA | 37  37 | .417  .72 | .14  -.06 |

Spearman correlations between change scores (baseline - 6-month follow-up) of clinical data and NMSS-T. Bold font highlights significant results, *p*<.05. Positive correlations indicate that higher changes in clinical scores are associated with higher changes in NMSS-T.

**Abbreviations:** LEDD: Levodopa Equivalent Daily Dose; LEDD-DA: LEDD of Dopamine Agonists; NMSS-T: Non-Motor Symptom Scale total score; PDQ-8 SI: = 8-item Parkinson’s Disease Questionnaire summary index; SCOPA = Scales for Outcome in Parkinson’s Disease.

**Image analysis**

**Table e-2 Association between FA-Values and NMSS-T**

| Positive  Cluster | Location | Slope | Intercept | p-Value | Volume in mm³ | MNI152-Coordinates | | |
| --- | --- | --- | --- | --- | --- | --- | --- | --- |
|  |  |  |  |  |  | X | Y | Z |
| P1 | Right insular cortex | .0002 | .1336 | .032 | 183 | 40 | -17 | 7 |
| Negative Cluster |  |  |  |  |  |  |  |  |
| N1 | Right cingulum | -.0004 | .4038 | < .001 | 436 | 13 | 28 | 31 |
| N2 | Left Heschl’s gyrus Left inferior longitudinal fasciculus | -.0006 | .3364 | < .001 | 282 | -40 | -26 | 5 |
| N3 | Left cingulum | -.0004 | .4528 | .014 | 203 | -13 | 28 | 18 |

**Table e-2.** Characteristics of clusters with an association between PD patients’ FA-values and postoperative change in NMSS-T. “Positive Cluster” denotes clusters with a positive association between patients’ FA-values and percentage differences in NMSS-T, i.e. higher FA-values were associated with higher postoperative values. “Negative Cluster” denotes clusters with a negative association between patients’ FA-values and percentage difference of NMSS-T, i.e. higher FA-values were associated with lower postoperative values. “Location” indicates the anatomical landmark comprising the majority of voxels of a cluster according to Johns Hopkins University (JHU) white matter atlas, Harvard-Oxford cortical and subcortical atlas, and University College London (UCL) cerebellar atlas. P-Values are clusterwise p-values corrected for multiple comparisons. “Volume in mm³” denotes the size of a cluster and “MNI152-coordinates” describes the coordinates of the cluster’s center of gravity in MNI152-space.

**Table e-3 Association between ODI-Values and NMSS-T**

| Positive  Cluster | Location | Slope | Intercept | p-Value | Volume in mm³ | MNI152-Coordinates | | |
| --- | --- | --- | --- | --- | --- | --- | --- | --- |
|  |  |  |  |  |  | X | Y | Z |
| P1 | Right putamen Right superior longitudinal fasciculus | .0003 | .3574 | < .001 | 223 | 31 | -6 | 7 |
| P2 | Right precuneous cortex Right cingulate gyrus (posterior division) | .0008 | .2567 | .008 | 159 | 6 | -36 | 43 |
| P3 | Right putamen Right anterior thalamic radiation | .0004 | .3853 | .024 | 141 | 19 | 13 | -2 |
| P4 | Left forceps major  Left cuneal cortex Left intracalcarine cortex | .0003 | .2911 | .034 | 135 | -11 | -79 | 18 |
| Negative Cluster |  |  |  |  |  |  |  |  |
| N1 | Left corticospinal tract | -.0002 | .1987 | < .001 | 785 | -25 | -17 | 29 |
| N2 | Right corticospinal tract | -.0002 | .2191 | < .001 | 306 | 23 | -10 | 25 |
| N3 | Left superior longitudinal fasciculus  Left superior parietal lobule | -.0002 | .2007 | .038 | 134 | -28 | -40 | 37 |
| N4 | Left occipital fusiform gyrus | -.0003 | .3549 | .047 | 130 | -15 | -74 | -15 |

**Table e-3.** Characteristics of clusters with an association between PD patients’ ODI-values and postoperative change in NMSS-T. “Positive Cluster” denotes clusters with a positive association between patients’ ODI-values and percentage differences in NMSS-T, i.e. higher ODI-values were associated with higher postoperative values. “Negative Cluster” denotes clusters with a negative association between patients’ ODI-values and percentage difference of NMSS-T, i.e. higher ODI-values were associated with lower postoperative values. “Location” indicates the anatomical landmark comprising the majority of voxels of a cluster according to Johns Hopkins University (JHU) white matter atlas, Harvard-Oxford cortical and subcortical atlas, and University College London (UCL) cerebellar atlas. P-Values are clusterwise p-values corrected for multiple comparisons. “Volume in mm³” denotes the size of a cluster and “MNI152-coordinates” describes the coordinates of the cluster’s center of gravity in MNI152-space.

**Table e-4 Association between NDI-Values and NMSS-T**

| Negative  Cluster | Location | Slope | Intercept | p-Value | Volume in mm³ | MNI152-Coordinates | | |
| --- | --- | --- | --- | --- | --- | --- | --- | --- |
|  |  |  |  |  |  | X | Y | Z |
| N1 | Left postcentral gyrus Left superior longitudinal fasciculus | -.0004 | .5663 | < .001 | 273 | -33 | -31 | 47 |
| N2 | Left cingulum | -.0003 | .5228 | .009 | 157 | -7 | -25 | 20 |
| N3 | Right forceps minor | -.0006 | .4678 | .049 | 125 | 12 | 19 | 15 |

**Table e-4.** Characteristics of clusters with an association between PD patients’ NDI-values and postoperative change in NMSS-T. “Negative Cluster” denotes clusters with a negative association between patients’ NDI-values and percentage difference of NMSS-T, i.e. higher NDI-values were associated with lower postoperative values. “Location” indicates the anatomical landmark comprising the majority of voxels of a cluster according to Johns Hopkins University (JHU) white matter atlas, Harvard-Oxford cortical and subcortical atlas, and University College London (UCL) cerebellar atlas. P-Values are clusterwise p-values corrected for multiple comparisons. “Volume in mm³” denotes the size of a cluster and “MNI152-coordinates” describes the coordinates of the cluster’s center of gravity in MNI152-space.

**Domain specific Statistics**

**Table e-5 Association between microstructural metrics and sleep/fatigue-outcomes**

| **Fractional Anisotropy** | | |  |  |  |  | | |
| --- | --- | --- | --- | --- | --- | --- | --- | --- |
| Positive  Cluster | Location | Slope | Intercept | p-Value | Volume in mm³ | MNI152-Coordinates | | |
|  |  |  |  |  |  | X | Y | Z |
| P1 | Right superior longitudinal fasciculus | .0003 | .3536 | .001 | 269 | 27 | -40 | 32 |
| P2 | Right postcentral gyrus | .0002 | .1475 | .005 | 233 | 55 | -19 | 21 |
| Negative Cluster |  |  |  |  |  |  |  |  |
| N1 | Left inferior longitudinal fasciculus  Left temporal fusiform cortex | -.0003 | .2373 | .02 | 194 | -41 | -38 | -12 |
| N2 | Right parietal operculum cortex | -.0002 | .3547 | .031 | 184 | 32 | -31 | 20 |
| **Orientation Dispersion Index** | | |  |  |  |  |  |  |
| Positive Cluster |  |  |  |  |  |  |  |  |
| P1 | Right corticospinal tract | .0001 | .0858 | < .001 | 297 | 24 | -17 | 8 |
| P2 | Right superior longitudinal fasciculus Right corticospinal tract | .0002 | .2448 | < .001 | 297 | 35 | -5 | 25 |
| P3 | Left corticospinal tract | .0001 | .1810 | < .001 | 270 | -26 | -22 | 40 |
| P4 | Right corticospinal tract | .0001 | .1694 | < .001 | 224 | 24 | -19 | 29 |
| P5 | Right planum polare Right middle temporal gyrus | .0002 | .2597 | .002 | 181 | 46 | -12 | -16 |
| P6 | Left inferior fronto-occipital fasciculus Left inferior longitudinal fasciculus | .0001 | .1169 | .002 | 179 | -30 | -27 | 3 |
| P7 | Right parietal operculum cortex | .0001 | .2207 | .009 | 157 | 37 | -33 | 21 |
| Negative Cluster |  |  |  |  |  |  |  |  |
| N1 | Right precentral gyrus | -.0004 | .4102 | < .001 | 223 | 11 | -13 | 45 |
| N2 | Left cuneal cortex Left forceps major | -.0004 | .3312 | .001 | 189 | -11 | -79 | 18 |
| N3 | Right lateral occipital cortex | -.0004 | .2939 | .043 | 131 | 30 | -71 | 26 |
| **Neurite Density Index** | | |  |  |  |  |  |  |
| Positive Cluster |  |  |  |  |  |  |  |  |
| P1 | Right corticospinal tract | .0002 | .5713 | .004 | 166 | 26 | -20 | 25 |

**Table e-5.** Characteristics of clusters with an association between PD patients’ microstructural metrics and postoperative change in NMSS-Domain 2 (Sleep/Fatigue). “Positive Cluster” denotes clusters with a positive association between patients’ microstructural metrics and postoperative changes in NMSS-Domain 2, i.e. higher values of a specific metric were associated with higher postoperative values. “Negative Cluster” denotes clusters with a negative association between patients’ microstructural metrics and postoperative difference in NMSS-Domain 2, i.e. higher values of a specific metric were associated with lower postoperative values. “Location” indicates the anatomical landmark comprising the majority of voxels of a cluster according to Johns Hopkins University (JHU) white matter atlas, Harvard-Oxford cortical and subcortical atlas, and University College London (UCL) cerebellar atlas. P-Values are clusterwise p-values corrected for multiple comparisons. “Volume in mm³” denotes the size of a cluster and “MNI152-coordinates” describes the coordinates of the cluster’s center of gravity in MNI152-space.

**Table e-6 Association between microstructural metrics and attention/memory-outcomes**

| **Fractional Anisotropy** | | |  |  |  |  | | |
| --- | --- | --- | --- | --- | --- | --- | --- | --- |
| Positive  Cluster | Location | Slope | Intercept | p-Value | Volume in mm³ | MNI152-Coordinates | | |
|  |  |  |  |  |  | X | Y | Z |
| P1 | Left anterior thalamic radiation | .0002 | .3534 | < .001 | 319 | -26 | 26 | 5 |
| P2 | Right precentral gyrus Right cingulate gyrus | .00017 | .1418 | .01 | 211 | 13 | -18 | 37 |
| P3 | Left insular cortex Left inferior fronto-occipital fasciculus | .00018 | .45 | .012 | 205 | -30 | -22 | -2 |
| P4 | Left cingulum | .00018 | .3651 | .031 | 185 | -17 | 24 | 27 |
| Negative Cluster |  |  |  |  |  |  |  |  |
| N1 | Right superior longitudinal fasciculus | -.00021 | .4261 | .006 | 225 | 33 | 7 | 20 |
| **Orientation Dispersion Index** | | |  |  |  |  |  |  |
| Positive Cluster |  |  |  |  |  |  |  |  |
| P1 | Right frontal pole | .0002 | .2707 | < .001 | 203 | 30 | 37 | 6 |
| P2 | Left parahippocampal gyrus  Left inferior fronto-occipital fasciculus | .0002 | .1944 | .049 | 128 | -27 | -7 | -10 |
| Negative Cluster |  |  |  |  |  |  |  |  |
| N1 | Right Putamen Right cerebral white matter | -.0002 | .2699 | < .001 | 1782 | 35 | -24 | 2 |
| N2 | Left Putamen Left cerebral  white matter | -.0003 | .2902 | < .001 | 916 | -29 | -18 | -1 |
| N3 | Left anterior thalamic radiation | -.0002 | .2264 | < .001 | 734 | -23 | 32 | 7 |
| N4 | Right corticospinal tract | -.0001 | .2051 | < .001 | 368 | 18 | -16 | 31 |
| N5 | Left cingulum | -.0002 | .2440 | < .001 | 267 | -16 | 23 | 30 |
| N6 | Left cerebral white matter | -.0002 | .2181 | < .001 | 248 | -16 | -5 | 37 |
| N7 | Left corticospinal tract | -.0001 | .1990 | .002 | 187 | -28 | -23 | 38 |
| N8 | Right anterior thalamic radiation Right Putamen | -.0002 | .4027 | .011 | 153 | 26 | 9 | 3 |
| N9 | Left anterior thalamic radiation Left cerebral white matter | -.0002 | .2531 | .023 | 141 | -5 | -3 | -9 |
| N10 | Left cerebral white matter | -.0002 | .2233 | .039 | 133 | -16 | 13 | 37 |
| **Neurite Density Index** | | |  |  |  |  |  |  |
| Negative Cluster |  |  |  |  |  |  |  |  |
| N1 | Right Putamen | -.0004 | .6042 | < .001 | 1380 | 31 | -18 | 6 |
| N2 | Right precentral gyrus Right superior longitudinal fasciculus | -.0003 | .6311 | < .001 | 450 | 28 | -12 | 41 |
| N3 | Left Putamen | -.0006 | .6524 | < .001 | 234 | -23 | -8 | 7 |
| N4 | Right corticospinal tract | -.0002 | .6063 | < .001 | 232 | 25 | -14 | 16 |
| N5 | Right corticospinal tract | -.0003 | .6590 | .009 | 152 | 15 | -32 | -26 |
| N6 | Left corticospinal tract | -.0004 | .6523 | .023 | 138 | 1 | -31 | -22 |
| N7 | Right V | -.0003 | .6293 | .031 | 133 | 6 | -61 | -22 |

**Table e-6.** Characteristics of clusters with an association between PD patients’ microstructural metrics and postoperative change in NMSS-Domain 5 (Attention/Memory). “Positive Cluster” denotes clusters with a positive association between patients’ microstructural metrics and postoperative changes in NMSS-Domain 5, i.e. higher values of a specific metric were associated with higher postoperative values. “Negative Cluster” denotes clusters with a negative association between patients’ microstructural metrics and postoperative difference in NMSS-Domain 5, i.e. higher values of a specific metric were associated with lower postoperative values. “Location” indicates the anatomical landmark comprising the majority of voxels of a cluster according to Johns Hopkins University (JHU) white matter atlas, Harvard-Oxford cortical and subcortical atlas, and University College London (UCL) cerebellar atlas. P-Values are clusterwise p-values corrected for multiple comparisons. “Volume in mm³” denotes the size of a cluster and “MNI152-coordinates” describes the coordinates of the cluster’s center of gravity in MNI152-space.

**Table e-7 Association between microstructural metrics and urinary-outcomes**

| **Fractional Anisotropy** | | |  |  |  |  | | |
| --- | --- | --- | --- | --- | --- | --- | --- | --- |
| Positive  Cluster | Location | Slope | Intercept | p-Value | Volume in mm³ | MNI152-Coordinates | | |
|  |  |  |  |  |  | X | Y | Z |
| P1 | Right cingulate gyrus, anterior division | .0003 | .3113 | < .001 | 273 | 8 | 19 | 26 |
| P2 | Left superior longitudinal fasciculus | .0003 | .1206 | .01 | 209 | -43 | -37 | 22 |
| Negative Cluster |  |  |  |  |  |  |  |  |
| N1 | Left inferior fronto-occipital fasciculus | -.0004 | .2375 | .004 | 235 | -32 | 32 | 3 |
| **Orientation Dispersion Index** | | |  |  |  |  |  |  |
| Positive Cluster |  |  |  |  |  |  |  |  |
| P1 | Left Pallidum  Left Putamen Left anterior thalamic radiation | .0003 | .2993 | < .001 | 198 | -16 | 2 | -6 |
| P2 | Left VI | .0003 | .3427 | .005 | 167 | -17 | -70 | -18 |
| P3 | Left V | .0004 | .2985 | .031 | 137 | -11 | -60 | -10 |
| **Neurite Density Index** | | |  |  |  |  |  |  |
| Positive Cluster |  |  |  |  |  |  |  |  |
| P1 | Left Pallidum  Left Putamen Left anterior thalamic radiation | .0006 | .5718 | < .001 | 226 | -15 | 2 | -7 |
| P2 | Left VI | .0005 | .4453 | < .001 | 193 | -13 | -75 | -15 |
| P3 | Right Pallidum  Right Putamen | .0003 | .5266 | .002 | 177 | 12 | 10 | -10 |
| P4 | Left V | .0006 | .4012 | .002 | 172 | -5 | -62 | -9 |

**Table e-7.** Characteristics of clusters with an association between PD patients’ microstructural metrics and postoperative change in NMSS-Domain 7 (Urinary). “Positive Cluster” denotes clusters with a positive association between patients’ microstructural metrics and postoperative changes in NMSS-Domain 7, i.e. higher values of a specific metric were associated with higher postoperative values. “Negative Cluster” denotes clusters with a negative association between patients’ microstructural metrics and postoperative difference in NMSS-Domain 7, i.e. higher values of a specific metric were associated with lower postoperative values. “Location” indicates the anatomical landmark comprising the majority of voxels of a cluster according to Johns Hopkins University (JHU) white matter atlas, Harvard-Oxford cortical and subcortical atlas, and University College London (UCL) cerebellar atlas. P-Values are clusterwise p-values corrected for multiple comparisons. “Volume in mm³” denotes the size of a cluster and “MNI152-coordinates” describes the coordinates of the cluster’s center of gravity in MNI152-space.

**Supplementary Figures**

**Figure e-1.**

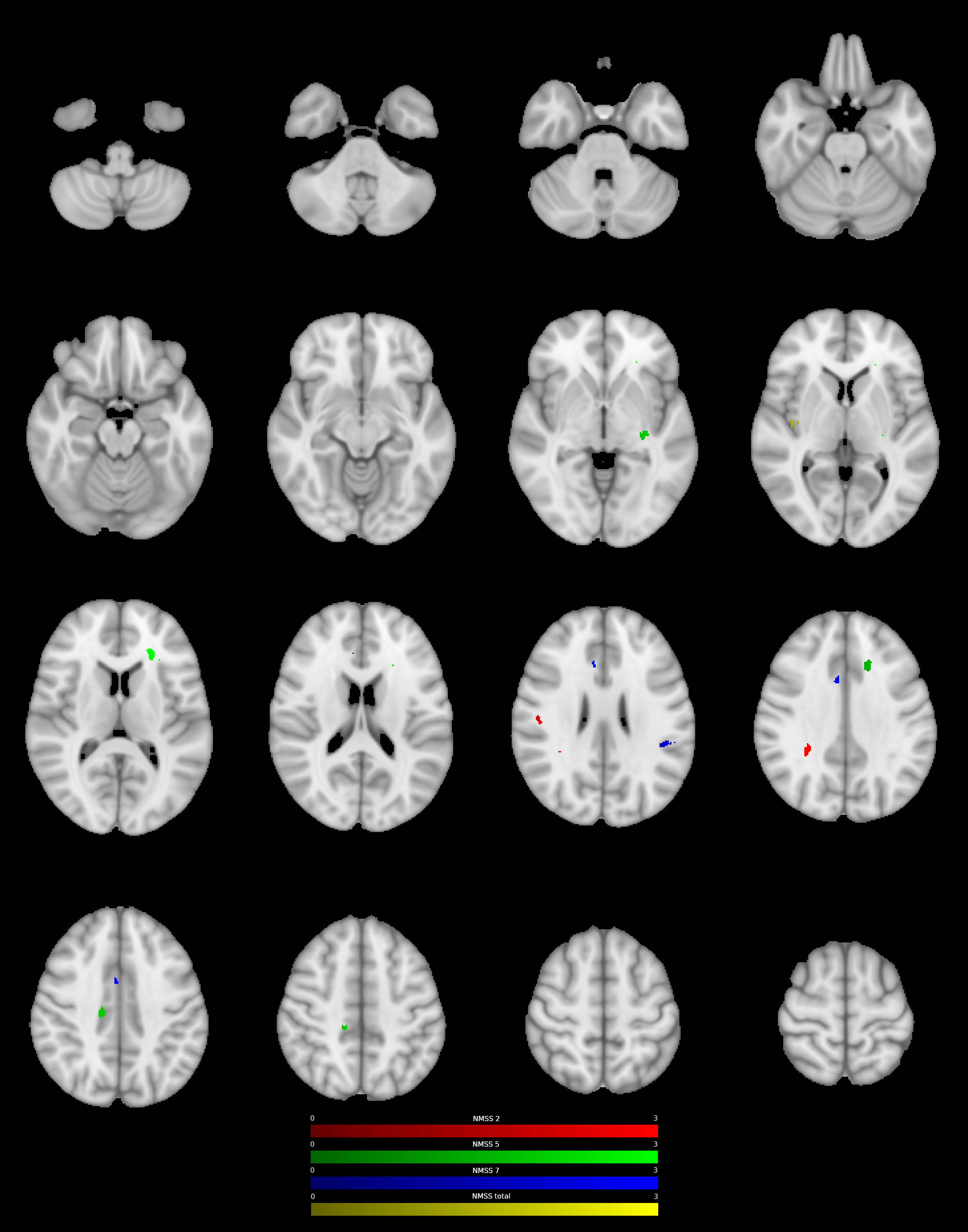

**Figure e-1.** Clusters with a positive association between PD patients’ FA-values and postoperative change in NMSS-T (yellow), Domain 2 (sleep/fatigue, red), Domain 5 (attention/memory, green), and Domain 7 (urinary, blue), as revealed by the whole brain analysis. P-Values were corrected for multiple comparisons using a permutation-based approach.

**Figure e-2**

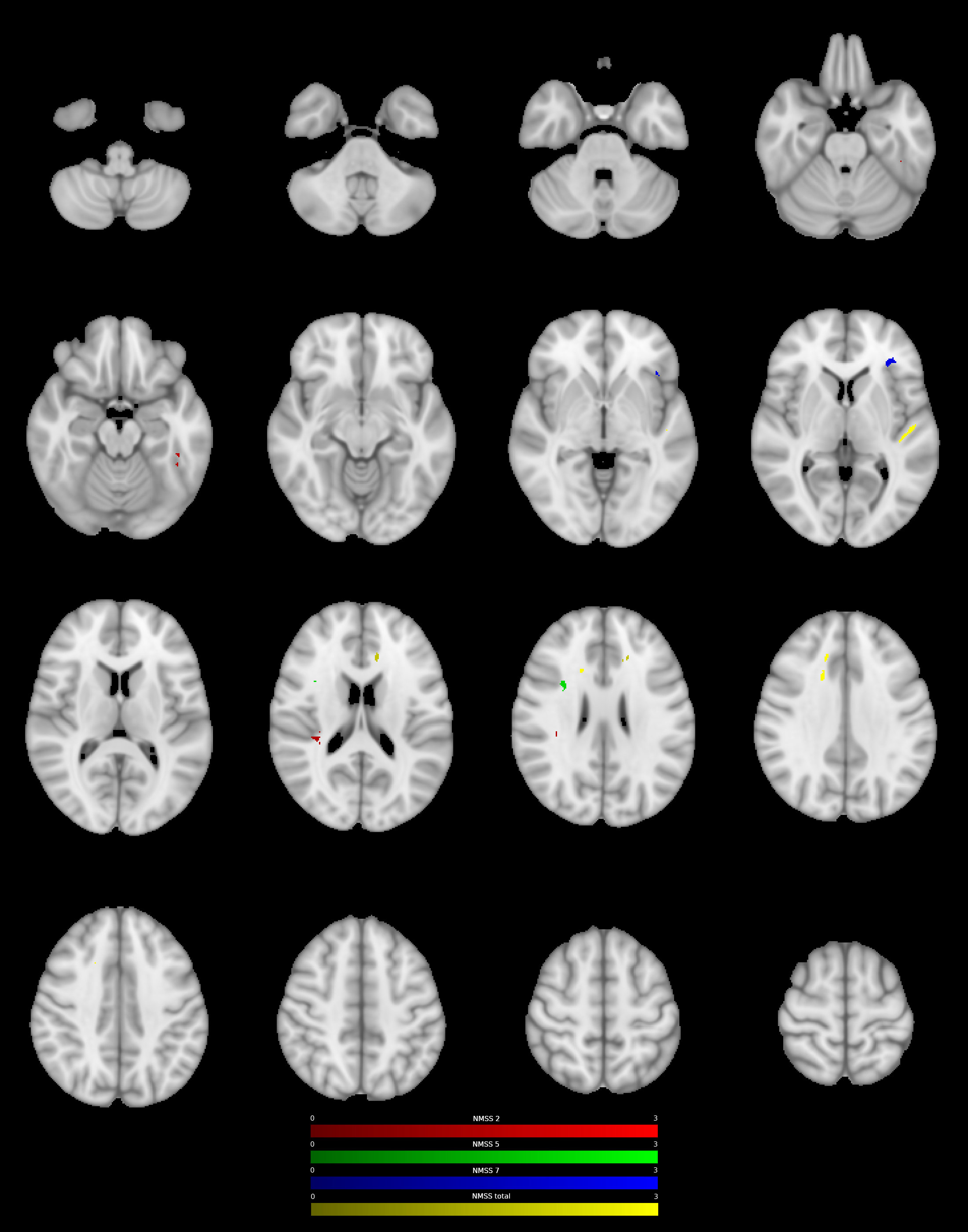

**Figure e-2.** Clusters with a negative association between PD patients’ FA-values and postoperative change in NMSS-T (yellow), Domain 2 (sleep/fatigue, red), Domain 5 (attention/memory, green), and Domain 7 (urinary, blue), as revealed by the whole brain analysis. P-Values were corrected for multiple comparisons using a permutation-based approach.

**Figure e-3**

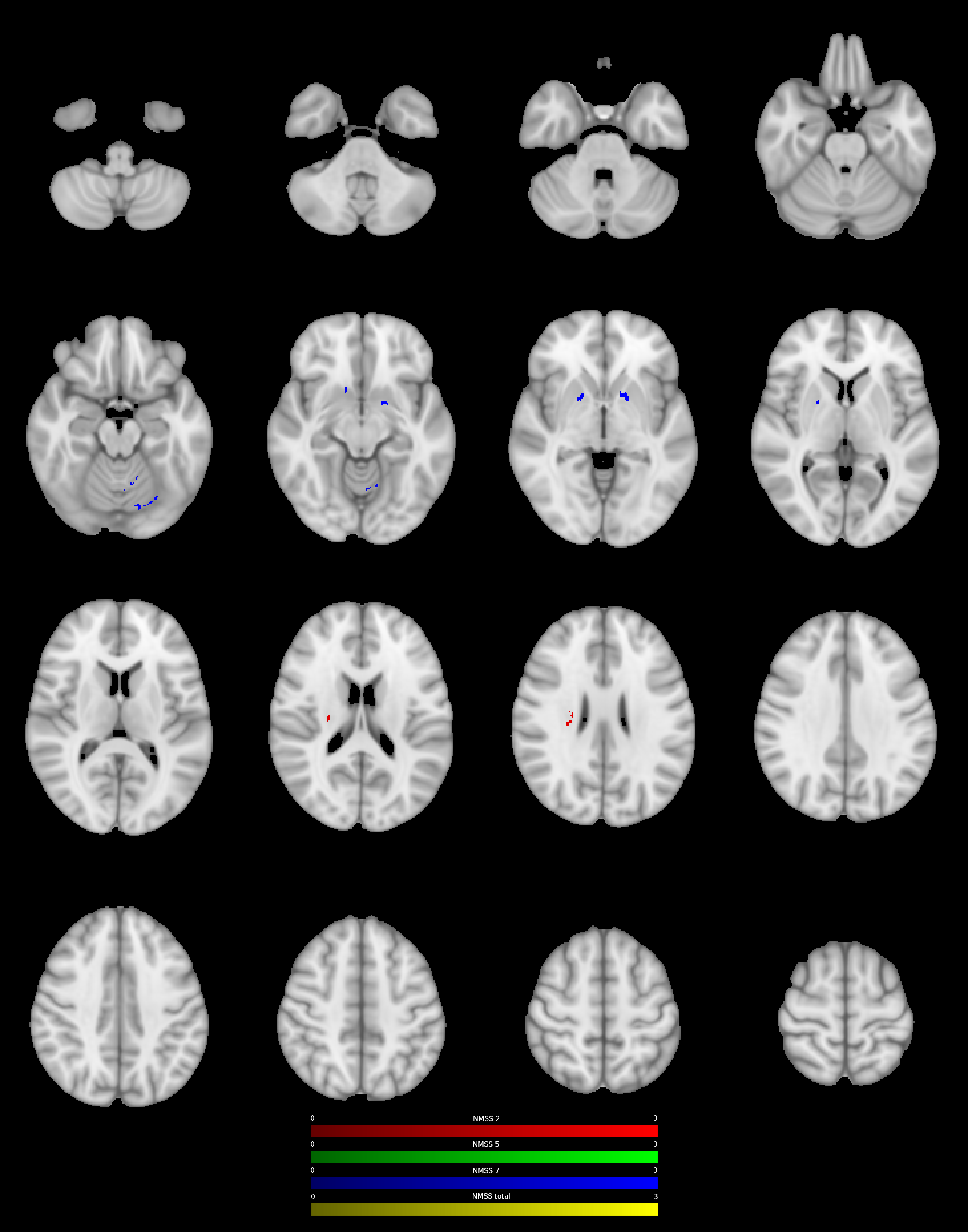

**Figure e-3.** Clusters with a positive association between PD patients’ NDI-values and postoperative change in NMSS-T (yellow), Domain 2 (sleep/fatigue, red), Domain 5 (attention/memory, green), and Domain 7 (urinary, blue), as revealed by the whole brain analysis. P-Values were corrected for multiple comparisons using a permutation-based approach.

**Figure e-4**

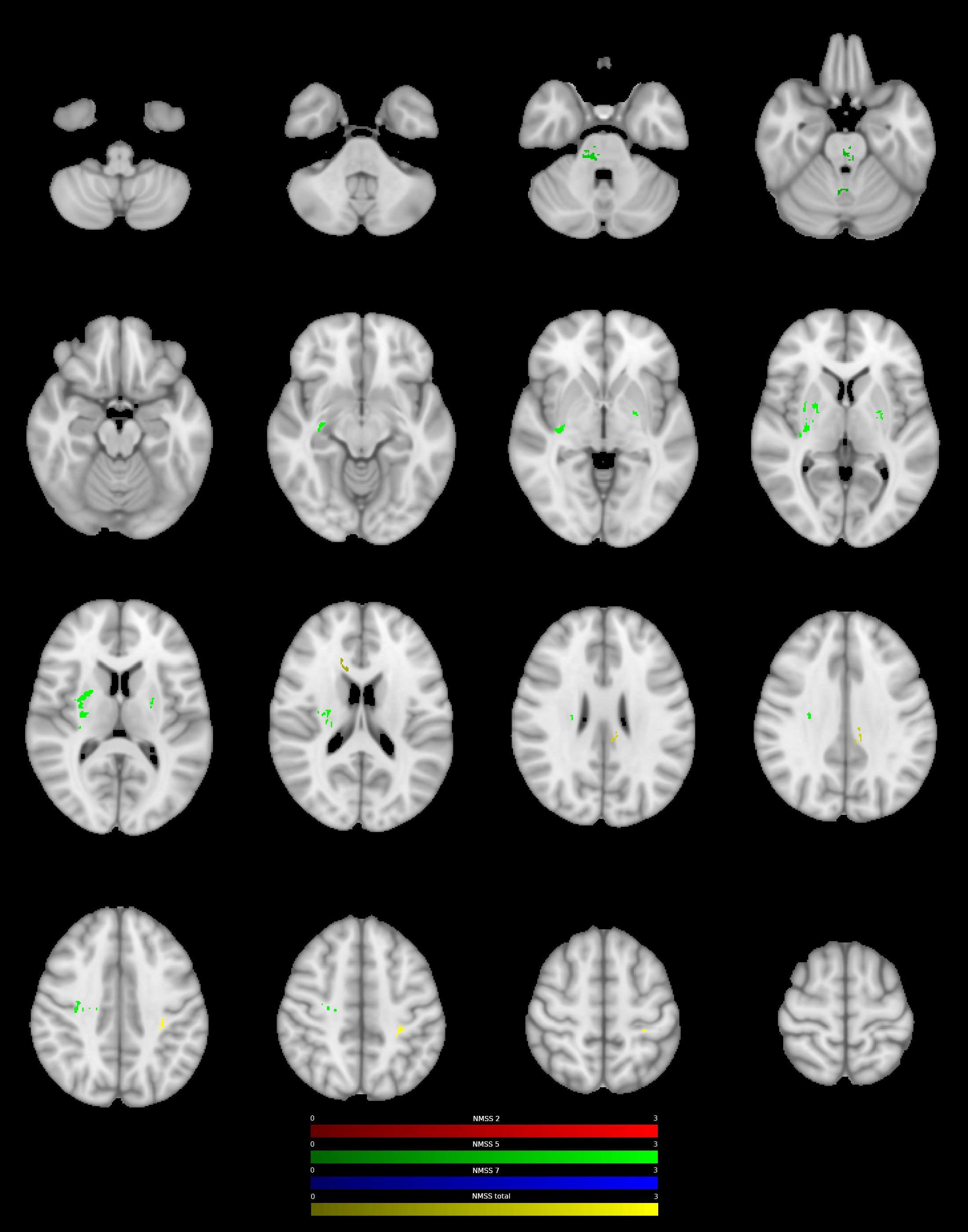

**Figure e-4.** Clusters with a negative association between PD patients’ NDI-values and postoperative change in NMSS-T (yellow), Domain 2 (sleep/fatigue, red), Domain 5 (attention/memory, green), and Domain 7 (urinary, blue), as revealed by the whole brain analysis. P-Values were corrected for multiple comparisons using a permutation-based approach.
